# Public perspectives on the trade-off between concentrating and dispersing health care benefits

**DOI:** 10.64898/2026.09.11.26362820

**Authors:** Laura Trigg, Ninian Schmeising-Barnes, Caroline Dix, Colin Angus, Aki Tsuchiya

## Abstract

**Background:** Conventional cost-effectiveness analysis assumes societal indifference to the distribution of health gains across potential patients, yet evidence suggests public preferences vary between concentrating and dispersing benefits, depending on the size of health gain. The handful of Person Trade-Off (PTO) studies that explored this did not examine the cognitive mechanisms and rationales underlying the PTO tasks.

**Objectives:** To qualitatively investigate how individuals conceptualise and justify trade-off decisions within a PTO task designed to elicit distributional preferences regarding healthcare benefits.

**Methods:** Following a pilot phase (n=10), online cognitive debrief interviews (n=10) were conducted with UK adults using a think-aloud protocol and concurrent verbal probing. Participants completed an interactive, web-based PTO task that presented choices between concentrating larger health gains among fewer recipients or dispersing smaller gains across a larger population. Data were analysed using codebook thematic analysis.

**Results:** Six primary themes emerged across two categories: decision-making processes and decision drivers. Participants displayed varying degrees of moral discomfort, utilizing cognitive mechanisms ranging from strict mathematical logic (QALY maximisation) to emotion-driven heuristics. Key decision drivers included patient age, quality of survival, societal productivity, and strong death aversion. Participants frequently rejected pure QALY maximisation and held strong egalitarian preferences to give every patient a chance to receive treatment.

**Conclusion:** Public perspectives on benefit distribution are highly heterogeneous and shaped by complex normative values rather than pure health maximisation. Understanding these qualitative rationales is essential for isolating genuine distributional preferences from cognitive shortcuts, aiding health technology assessment bodies in evaluating equity weights.

## Introduction

In the UK, healthcare reimbursement decisions are made based on cost-effectiveness, where effectiveness is measured in terms of quality adjusted life years (QALYs) gained. QALYs are a measure combining both quality and quantity of life. In most cases, all QALYs are valued by society equally regardless of the recipient. Thus, when the evaluation aims to maximise health, it is on the sole basis of QALY gains, irrespective of recipient (1). However, evidence suggests that this does not always hold; health gains may be valued differently depending on how they are distributed across the population, or on the characteristics of the recipients. For example, value may vary depending on patient attributes such as age, presence of dependents, or initial illness severity (1–3). Beyond individual characteristics, societal preferences also shape how health gains are distributed (4).

Preferences for dispersion mean that for a specific level of total health gain, society prefers to share health gains across more recipients, with each recipient receiving less. This means that each additional QALY given to the same individual is valued less than the previous one. Conversely, preference for concentration is where society places greater value on larger benefits being allocated to fewer individuals overall, and each additional QALY given to the same individual is valued more than the last. Preferences for concentration and dispersion may differ based on the size of the health gain per patient; prior studies (5,6) have found that when per-person health gains are small, participants prefer concentration; when gains are large, they prefer dispersion, creating an S-Shaped curve. When visualising the relationship between the health gain per person and associated social value, concentration preferences were plotted as a convex curve, and dispersion preferences as concave (Figure 1).

**Figure 1:**
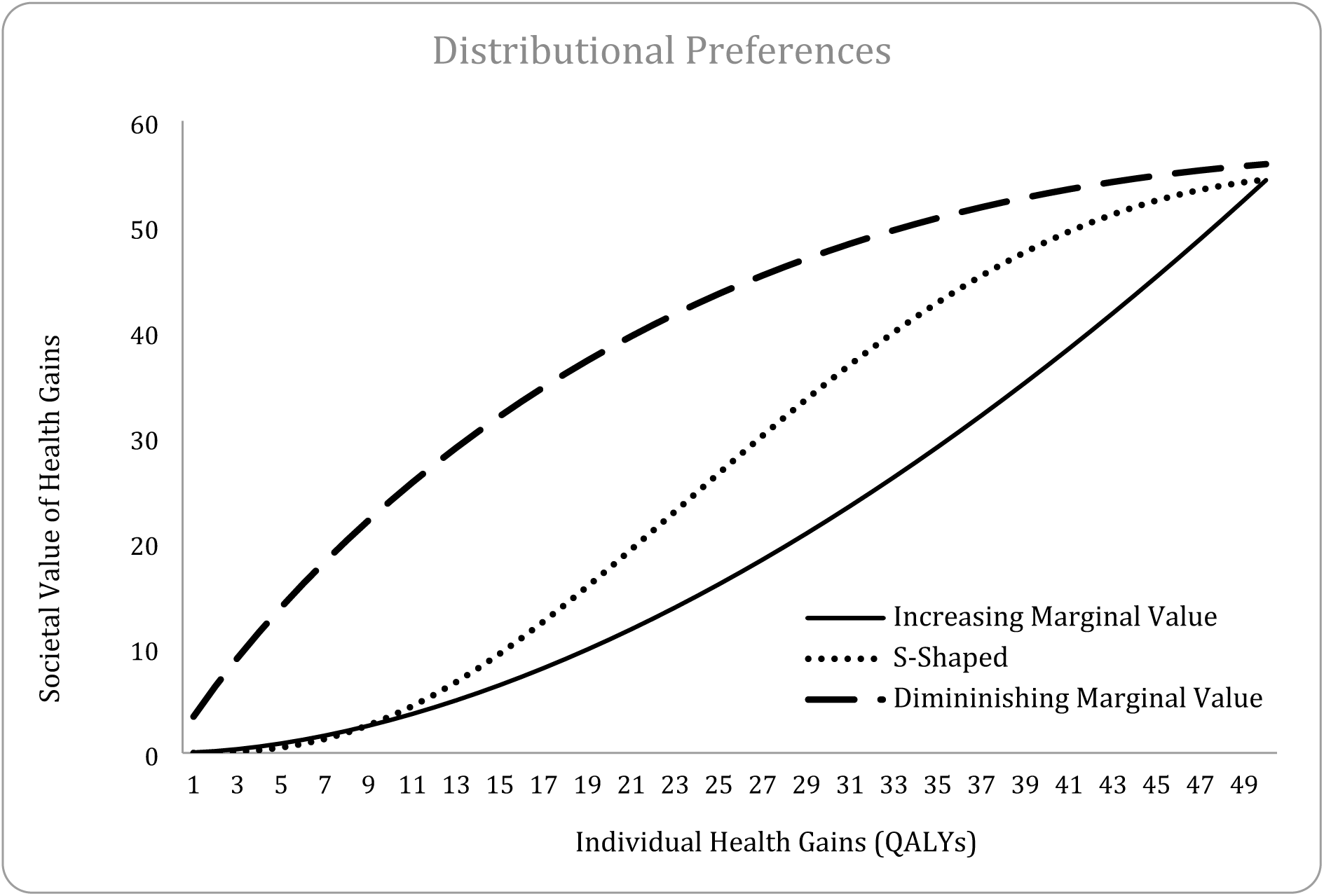
Distributional Preferences (6).

These studies, however, have assumed that any health gain occurs only in the form of a life extension without specifying to the participants the quality of life of the patients. This allows for confounding preferences around concentration and dispersion with severity preferences as participants may make inferences about the participants’ level of health based on the additional time they may live for. QALYs may be gained through improving quality of life *or* extending life span, and distributional preferences may depend on the starting health level (7). Additionally, previous studies did not specify what happens to the patients not receiving treatment. The lack of information on the counterfactual may have also influenced their preferences by drawing focus to patients who are going to receive the treatment rather than those who are not.

From past and current guidance in the UK and Internationally (particularly in Norway and The Netherlands) (8,9), preferences have been incorporated into health economic decisions by using a QALY weighting (10,11), but currently any additional QALY weights relating to equity are not considered due to a lack of consensus (12). However, if equity-related weights are to be considered in future guidance, it is important to understand why certain preferences exist.

Although quantitative preference tasks reveal non-linear curves, they cannot isolate true distributional preferences from cognitive shortcuts. Investigating the qualitative rationale behind choices is critical to assess task comprehension and ensure the validity of health state valuations. In addition, data gathered from a cognitive debrief exercise can provide insight into how individuals conceptualise the trade-offs, the values that underpin their preferences and the attributes of the questions that are prioritised in decision making. Quantitative research on distributional preferences is already sparse, and a qualitative investigation into understanding the rationale behind these preferences has not been carried out before. This study will explore preferences around the concentration and dispersion of healthcare. The aim of this paper is to understand and examine how individuals make trade-off decisions in a Person Trade-Off (PTO) task designed to elicit distributional preferences.

## Methods

There were three phases to this study; a pilot, cognitive debrief interviews, and a quantitative survey. All stages used the same (PTO) tasks. This paper reports on the pilot study and cognitive debrief interventions.

Ethical approval from this research was obtained from The University of Sheffield (066410). All participants were compensated £25 for their time.

### Person Trade-off Task

The PTO task was adapted from an earlier study (6). It was developed and administered using R-Shiny - a web-based app, considering the conceptual issues raised in a systematic literature review (13) and by Peasgood et al (14). It presented participants with a hypothetical population, consisting of 200 patients aged 20-year-old, with the same Health Related Quality of Life (HRQoL) Score. The participant was asked to imagine they were the decision maker responsible for deciding how the healthcare was allocated across this population. The level of health was presented to the participants as a proportion, 100% represented full health, and 0% represented being dead. The HRQoL scores used in the question were 20%, 40%, 60%, 80% and 100%. Participants were told that if the hypothetical patient did not receive the treatment, they would die immediately, and if they did receive the treatment, they would live for the specified number of years in the same health state and then die. The participant was then shown two scenarios, A and B. In scenario A, participants were told how many patients would receive the treatment, and how long they would live for after they received it. In scenario B, any patient who received the treatment would live for 10 years. The participant was asked to determine how many patients should receive the treatment in scenario B to make it ‘feel just as good as’ scenario A. To illustrate the scenario, the survey used interactive visual aids.

Dynamic boxes containing 200 stick figures were presented for each scenario, where those receiving the treatment were depicted in black, and those not receiving it in grey. These visuals updated in real-time as participants entered their responses. Examples of these questions are found in Appendix A.

### Procedure

The pilot phase aimed to collect feedback on the way participants understood PTO tasks and to build on them. The cognitive debrief phase following the improvements aimed to gain deeper insights into the rationale behind the way participants responded to the PTO tasks. There were no further changes made based on feedback from this stage.

For both the pilot and cognitive interviews, participants received an information sheet and consent form (returned on google forms). Once consent was given, participants were invited to book an interview slot. All interviews were conducted online via Google Meets in a think-aloud style (15), where participants were asked to complete the survey whilst sharing their screen with the interviewer and asked to verbalise their thoughts during completion. In most cases, both parties had their camera on. During the task, concurrent verbal probes were used to encourage the participant to expand or reflect upon their answers (16). Once the task was completed, participants were asked around 15 semi-structured interview questions (a full topic guide is available in Appendix B).

The lead researcher (LT) conducted all interviews. Participants were introduced to the study and think-aloud process, sent the link to the survey platform and were then asked to share their screen. For both stages, the PTO task was expected to take 20 minutes, with 40 minutes for the semi-structured interviews. The interviews were video recorded and transcribed verbatim using Google Meets. The recordings were used to check transcriptions manually (LT) and then deleted. Transcriptions were anonymised and are available upon request.

### Recruitment

There were 10 participants recruited for the pilot phase. For the cognitive debrief interviews, information power was assessed after 10 participants had completed interviews. This assessment involved a review of the transcripts against Malterud et al’s (17) dimensions of information power. Given the narrow scope of the interview questions and the strong quality of dialogue, it was determined that sufficient information power was achieved and study objectives were able to be achieved without further recruitment. Any participants who participated in the pilot study were excluded from the cognitive debrief. All participants were recruited through Prolific Academic, were ≥ 18 years old, a UK resident and self-reported to be proficient in English.

### Cognitive Debrief Data Analysis

There was no formal data analysis conducted at the pilot phase. For the cognitive debrief interviews, the data were analysed using codebook thematic analysis, a method seen as a ‘middle ground’ between coding reliability approaches and reflexive thematic analysis (18,19). This approach allowed for a structured, team-based analysis whilst maintaining a data-driven focus.

Coding was done by inductive analysis and refinement through team-based consensus. A positionality statement can be found in Appendix C.

Firstly, the lead researcher (LT) inductively coded all 10 transcripts from the cognitive debrief interviews. Based on this initial analysis, a codebook was developed, containing the code names, descriptions and examples. To enhance the rigour of the analysis and mitigate individual bias, two co-authors (CD, NSB) served as secondary coders. Using the codebook, CD and NSB independently coded a subset of the transcripts (5 each).

Following coding, the three coders held consensus meetings to compare coding applications through negotiated agreement, discussing discrepancies until a consensus was reached on the interpretation of the data. Validated codes were collated into candidate themes by the lead researcher, which were then reviewed and refined collaboratively by the coders to ensure they accurately captured the patterns within the dataset and provided a coherent narrative of the participants’ experiences.

## Results

### Pilot Interviews

Ten participants, aged between 25 – 56, completed the pilot interviews in September 2025. A table of design changes made can be found in Appendix D; some changes involved renaming Option A and B to Scenario A/B to emphasise that the participants are not required to choose between options. Additional example pages were added, and the pop-up response confirmation were made clearer. Initially, the participants were given a slider to indicate the number of patients in Scenario B that would make it feel just as good as Scenario A, with a maximum value of 20, but many commented that the maximum input in the slider was not large enough. To avoid any bias towards dispersion or concentration with respect to the maximum number of patients in the slider, this was replaced with a free text input box, allowing the maximum answer to be the size of the entire hypothetical population. Generally, the level of understandability of the task was high and participants commented that diagrams were helpful in the decision-making process. Mean completion time for the PTO task was approximately 20 minutes, though it varied between 15-45 minutes.

### Cognitive Debrief Interviews

Ten participants, aged between 24 and 57, participated in a cognitive debrief interview in October 2025. Five participants were in full-time employment, two were in part-time employment, one was not in paid work, and two did not disclose their employment status. Four of the participants identified as white, three as Black, one person each respectively identified as mixed or Asian, and one did not disclose.

Following the initial transcription of the first five interviews, the velocity of new code generation significantly decreased (20). Although the sampling criteria were relatively broad, the scope of the interviews was narrow, as they were guided by a structured PTO survey comprising ten fixed questions with limited variation. For most participants, the interviews were characterised by a high quality of dialogue and strong engagement, resulting in rich and in-depth data.

The data were coded using 41 distinct codes; quotes from the transcript were coded in larger sections to keep context and were frequently double coded to capture all codes within each quote.

Overall, there were six themes across the codes, categorised into two; the first set of themes fall under ‘decision making processes’, which focuses on how the participants made the decisions in the PTO questions, and the degree to which they struggled to do so. The second category of themes was ‘decision drivers’ which focuses on what attributes of the PTO the participants focused on, and the context in which the participants made the decisions.

### Themes

A thematic map showing the themes can be seen in Figure 2. There were two themes, in the yellow bubbles, that focused specifically on the decision-making process and four themes, in the green bubbles, that focused on the aspects of the question the participants valued. The theme ‘societal focus’ contains two sub-themes; societal responsibility and giving everyone a chance.

**Figure 2:**
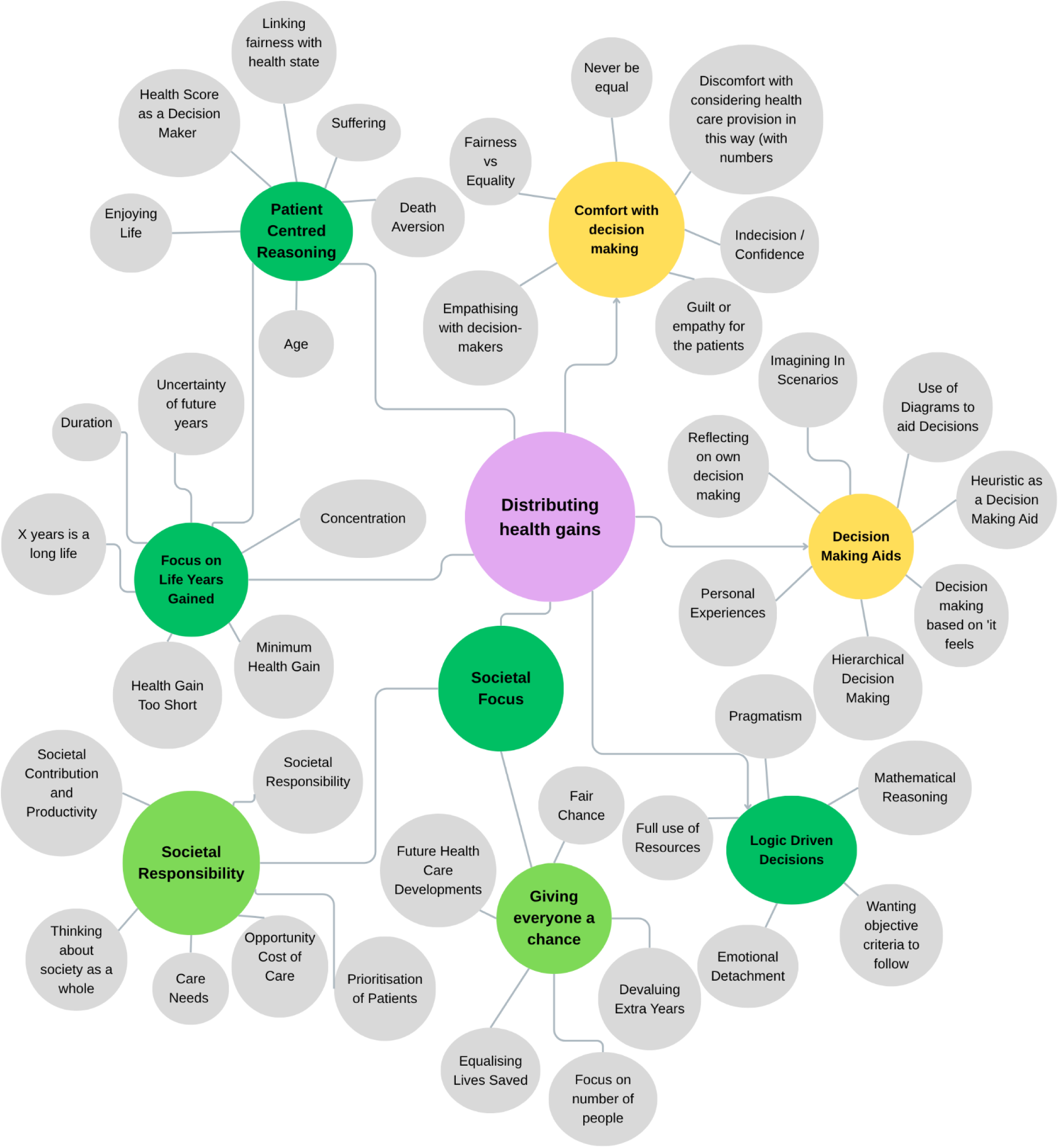
Themes and Codes.

#### Theme 1: Comfort with the decision-making

This theme captures the moral discomfort participants experienced when completing the elicitation task. Rather than viewing the scenarios as academic tasks, participants reflected on the tangible consequences of their choices, resulting in a sense of burden that extended beyond the immediate survey context. The act of decision-making in this context was often accompanied by feelings of guilt or empathy, both for the patients in the scenarios, and for those tasked with making these decisions in a real-world setting, noting, ‘they wouldn’t want to be put in that position’.

> *‘So it’s really sad to me. I’m like wow. damn, how do we do it to make things equal? I see why people who are in higher authorities especially when it comes to determining how we share our natural resource or in the medical line amongst people that’s why it’s always a tough decision for them but most times people would just feel they’re not doing enough, they’re not doing enough but whereas they’re doing a lot. Imagine I’m placed in this kind of scenario like real life and what am I supposed to do you know it’s not something you have to rush and provide an answer for.’* [P3]

The discomfort sometimes stemmed from the decision-making process itself, with some participants expressing unease with reducing health-care decisions to ‘pure numbers’. Consequently, some participants expressed that the two scenarios ‘will never be equal’; this was not due to mathematical impossibility, but because trading-off actual lives would never result in any solution that felt fair:

> *‘I don’t ever think it is going to feel equal. It’s never going to feel equal. It’s just about my perspective on being fair.’* [P9]

As patients receiving treatment in Scenario B always lived for an additional 10 years, participants sometimes felt the difference between health gain across scenarios was too great to feel equal regardless of number of recipients (for example, in Scenario A 200 patients gained 6 months each). This may reflect a threshold effect, where patients must achieve a minimum viable health gain before population trade-offs can occur.

When reflecting on the task, some participants drew distinctions between the concepts of ‘fairness’ and ‘equality’. While equality was viewed as a measurable, outcome-based metric, fairness was viewed as more subjective. One participant described equality as ‘thinking with your head’ and fairness as ‘thinking with your heart’.

There was a high degree of variation in participants’ confidence in their own responses, from certainty of response to almost paralysed indecision. However, there was a pattern across participants; those applying QALY maximisation principles expressed a higher level of confidence in the decisions, possibly due to the presence of a mathematically ‘correct’ answer, whereas participants using an emotion-based approach experienced an internal battle between wanting to save the highest number of people possible in Scenario A, but also trying to remain true to the survey and make the two scenarios feel equal.

> *‘I was very confident. Because that’s why I didn’t rush to give my answers. I try to put myself in the scenario. I try to create a scene in my head and I did everything that way. So that’s it. So I think I was 100%.’* [P3]

> *‘There was no right or wrong answer. There was and I was always unsure about my choices and if I was to do it again and again and to do it every day, I’d still be unsure every time’* [P9]

#### Theme 2: Decision making aids

This theme discusses the cognitive mechanisms, practical tools and heuristic strategies participants employed to navigate the complexity of the allocation tasks. Participants did not rely on a single mode of reasoning, instead relying on multiple decision-making aids simultaneously, such as visual aids, internal consistency and personal projection (18) to justify their choices participants frequently utilised the provided visual aids to ‘test’ potential answers before committing to a response. However, this often served to confirm a ‘gut feeling’, rather than a purely calculated preference. This suggests that while participants used the diagrams to visualise different answers, the decision itself was driven by other decision-making aids, including an intrinsic ‘sense of rightness’.

> *‘It looks about right with regards to visuals to make it equal. but again, in my head, there’s still that thought that why couldn’t it be 100 people getting 10 years. it’s still again additional life to what they would have if they didn’t have the treatment.’* [P5]

Some participants established a decision rule or internal logic early on and continued to apply these rules across subsequent questions throughout the survey.

> *‘I suppose it has to come back to the reason that I have on all the other questions’.* [P8]

The use of such choice behaviour allowed participants to avoid re-evaluating the trade-offs in each task. This consistency may reflect one of two mechanisms. Firstly, participants may have used early questions to clarify their own decision logic and subsequently applied this logic more systematically, consistent with learning effects. Alternatively, participants may have relied on prior answers as a cognitive shortcut, exhibiting anchoring (19) or choice-history dependence (21) rather than preference-based deliberation. The first of these was seen more often in participants who employed logic-based decision rules, as potentially the ‘rules’ were more well-defined and generalisable across different questions than those participants who answered using emotion-driven logic.

Participants also displayed two distinct psychological strategies for handling the emotional weight of the decision throughout; participants either distanced themselves from the decision (18,22), or viewed the decision through the lens of personal experience (23). With respect to distancing, some participants attempted to reduce the stakes of the decision, specifically the feeling of ‘playing god’ by reframing the health gain variable. By mentally converting ‘life years gained’ into ‘monetary value’, these participants abstracted the human element, making the trade-off feel more transactional:

> *‘Yeah. In this context, fairness means for me, if more persons are able to get to access something and you want to reduce the number of persons to assess that same thing. It shouldn’t mean you should reduce the number of……. okay let me use money. if you need to share £100 for 100 persons and you say okay you want to reduce the number of persons to access this £100 to 50. It shouldn’t make you reduce the number of money to £50.’* [P10]

Conversely, other participants leaned into the human element by reflecting on personal experiences. Participants drawing on memories of bereavement tended to have a stronger anecdotal preference for dispersion. In particular, when participants reflected on family illness, the focus shifted to potential suffering, with one participant stating, ‘the kindest thing to do is for him to pass quickly because he’s in pain, so that was the kind of way of my thinking’. In these instances, the decision was not made based on the abstract numbers on the screen, but on a projected realisation where any extension of life, regardless of Health Related Quality of Life, was viewed through the lens of personal loss and illness.

The remaining four themes fall into a ‘decision drivers’ category, where participants are explicitly considering the aspects of the question that factored into their decision.

#### Theme 3: Humanising the patients and Narrative Construction

This theme focuses on how participants looked beyond the abstract numerical data to a constructed reality of the individuals within the scenarios. Participants did not merely calculate outcomes; they engaged in a process of humanisation; actively visualising the patients to assess the qualitative value of the life years being allocated and constructing narratives regarding the hypothetical patient’s lives. This cognitive shift from statistical units to perceived ‘real people’ significantly raised the moral stakes of the decision. The act of non-selection of certain patients in the scenario was reframed not as a passive allocation of health gains, but as an active harm. Participants expressed a form of death aversion, feeling that failing to save a specific group was akin to ‘killing’ the individuals they had visualised.

> *‘…when I’m making this decision it’s almost that I am taking away the opportunity of the rest of the people not to live that life…’.* [P8]

Central to some participants’ reasoning was an assessment of the quality of survival the patients would experience. Participants generally considered the health score and the patient’s ability to enjoy additional life years. This introduced an ethical tension among some participants; while the primary drive was to extend life, this was counterbalanced when the health score was very low and participants wanted to avoid patient suffering. Generally, participants started to consider suffering when the health score was 40% or less. No participant commented that any of the given health states were worse than being dead (which would be a misrepresentation of the health score). However, some were conflicted when dealing with scenarios where patients would live for a long time in poor health.

> *‘…and while you want to save people, do you want these people to suffer 10 years if there is literally no change* [in the health score]*?’.* [P1]

Here, logic shifted from maximising years, to minimising suffering.

> *‘is it fair to suffer all these 10 years, or you’d rather just enjoy one year because obviously for sure your health is not…you can’t do lots of things’* [P1]

Whilst some participants tended to disregard the health score because it remained constant across all scenarios, others linked fairness with the health state. These participants noted that *if* the health score had differed between the two scenarios, they would have taken the health score into consideration more

> *‘if scenario A has a different health score and scenario B has a different health score, it could change my answer but it* [the health score] *covers all patients so it doesn’t matter’* [P10]

The age of the patients in the scenarios was also a dominant factor. Without prompting from the interviewer, participants explicitly noted that their decisions were influenced by the patients’ ages and that their choices might have differed had the patients been older. This suggests that participants did not necessarily view health gains as independent of age; rather, they appeared to use age as a heuristic for the productivity of additional years of life, assigning greater or lesser value based on what they perceived could be achieved during those additional years.

> *‘it would be fairer if a 20-year-old got an extra 10 years as opposed to a 70-year-old got an extra five years when realistically a 20-year-old is probably going to go out and do more in them 10 years than a 70-year-old would do in five years, that makes sense’.* [P8]

#### Theme 4: Focus on life years gained

In this theme, whilst participants still used patient-centred reasoning where the focus was on the patient rather than society, there was a specific focus on increasing marginal value of survival compared to constant marginal value of survival, where the years gained either held linear or non-linear value to participants. Participants expressed preferences for concentration by acknowledging the value in fewer people living for longer periods of time.

> *‘to make it equal means I need to concentrate on 10 people and give them more treatment but they for sure will be twice longer than the Scenario A’* [Scenario A: 20 people live for 5 years each, 100% health score] [P1]

This highlights a rejection of a strict QALY maximisation approach, where the total QALYs gained is the sole driver of value, in favour of a distributionally sensitive preference. Additional years gained held increasing marginal social value, as the extra time allowed patients to achieve a lot within it.

> *‘because let’s face it, you can’t get a lot done in six months anyway. So whereas the five people who’ve got the extra 20 years, at least they’ve got some more life to go at and they can make more of it’* [P9]

However, this was not always the case; some expressed doubt about whether patients would actually survive the full period, particularly when health scores were low, and accordingly discounted the value of very long life extensions (24).

> *‘it doesn’t make sense because if the health is poor, they’re unlikely to live for another 50 years, aren’t they?’* [P6]

Closely linked to the preference for concentration was the recognition of a minimum meaningful level of health where gains were seen as ‘worth it’. Participants articulated a floor effect, often cited between six weeks and two years, below which the extension of life was viewed as having little, or even negative utility, which was generally independent of the HRQoL.

> *‘why should the life of 200 persons be extended for 6 months and then after 6 months they will all die again? It’s useless. So it’s not valuable at all, anything below one year is almost useless’* [P10]

Participants believed that shorter extension to life did not alter the fundamental trajectory of the patient’s mortality; ‘it’s an extra 6 months, it doesn’t change anything’. However, the rejection of short life extension was not merely based on efficiency, but also on potential for psychological harm, suggesting that subjecting a patient to the knowledge of their impending death within 6 months may cause ‘emotional trauma’. In this view, the additional health gain was not seen as an extension to life, but an elongation of the dying process. Conversely, other participants displayed tension between the perceived lack of value of short health gains and the principle of societal equality. They stated that if given 6 weeks, this would personally be insufficient, but would support an additional 6 weeks of life on a societal level to ensure ‘everybody to get that chance’. This highlights a critical dissonance: personally a short duration feels valueless, whereas an egalitarian view encourages smaller gains to be shared to avoid excluding anyone.

#### Theme 5: Societal focus

This theme marks distinct participant priorities, with a movement beyond the benefit to the individual patients and instead focused on the implications of their choice on society as a whole. This theme is characterised by two sub-themes; giving everyone a chance, and societal responsibility.

#### Subtheme: giving everyone a chance (to receive treatment)

Many participants prioritised ‘lives saved’ over ‘life years gained’. Participants frequently made decisions based on dispersion preferences, where additional life-years to the same patients diminished in value once a basic threshold was met.

> *‘20 years is too much for five people, why don’t we save more people and they stay for 10 years?’* [P3]

Another participant stated that a gain of five or 10 years ‘doesn’t’ make much difference provided that ‘more people could get the treatment’. This reasoning suggests a non-linear value of additional health gain: whilst the difference between life and death is large, the difference between living for a short or a long period of time after the decision is negligible when considering the ‘cost’ of excluding others from a chance to live; ‘it doesn’t matter for how long they will live, definitely there should be more people saved’. Some participants could be categorised as ‘headcounters’, where they completely discounted any difference in the number of years and focused solely on the number of recipients of the care.

> *‘So in Scenario A, 200 people will get six months of life each, which is equal and fair to Scenario B, [where] 200 people are going to get 10 years of life each.’* [P9]

These preferences were often rationalised through hope for future health care developments, where participants again thought beyond the information given in the survey in favour of a real-world view, arguing that keeping a patient alive now, even in a poor health state, buys time for future health care.

> *‘So for me it doesn’t matter how many years it’s more about how many people will be alive and maybe at some point within five years or 10 years maybe they’ll get something that might improve their health.’* [P1]Preferences were sometimes rationalised by the possibility of future healthcare developments; if there is hope of medical developments, a shorter health gain is not a final sentence, but a bridge to a time when ‘some treatment might pop-up’.

However, participants did not always consider how the duration of the gain might impact the likelihood of being alive when such developments might occur. Thus, such rationales appear less as a calculation of future health but more a justification for prioritising the number of people receiving treatment.

There was a central focus around giving everyone a chance to receive some health gain; with some participants defining fairness as giving ‘everybody equal opportunity’. Participants explicitly contrasted their survey choices with real-world inequalities, contrasting the hypothetical scenario of the survey with the real world.

> *‘Being fair to me is…being fair was giving everybody equal opportunity to do whatever or giving them a chance to live in this scenario. So that’s what being fair actually mean…not just giving some few people advantage over the other people like what we see in the world right now the rich folks gets in fact premium services than the random people.’* [P3]

#### Subtheme: Societal Responsibility and Productivity

Some participants made their decisions based on the needs of the society as a whole. Participants measured the value of the healthcare program by its potential productivity and removal of societal burden, rather than based on the value to the individual patients themselves.

> *[Participant asked how they balanced the needs of the individuals compared to the population] ‘I think more population than individual. It was seen more society and what was fair within society rather than the actual individual and the individuals that would have been living for that amount of time if that makes sense. Again, I feel especially with something like our healthcare service we got here it’s all about fairness and people feeling fairness and that sort of social contract we’ve got in place with regards to NHS and things like that. It was definitely looking at the things in a whole rather than each individual circumstance.’* [P5]

Health was sometimes conceptualised as a form of human social capital essential for societal progress. Participants justified their decisions by linking physical health directly to economic and societal productivity, noting that a ‘healthy society is a wealthy society’. Several different factors were considered, one participant focused on providing treatment to enough people to ensure they are able to create future generations.

> *‘25 people will live for 10 years and within that 10 years they’ll be able to regenerate and we’re going to have more than 50 people on earth after that’* [P3]

Another patient focused on the specific contributions of the individual patients to society;

> *‘it also comes into the thing of what these people would do with their lives…suppose the higher the number the more potential for getting people that have interests in research into cancer and autoimmune diseases and other diseases’* [Scenario A: 5 people living for 20 years, 80% health] [P4]

Here, the rationalisation of decisions was driven not by the impact to the individual but based on what would result in the best outcome for the society.

Illness was sometimes framed as a ‘toll’ or ‘liability’. There was a tension for some participants between wanting to look after as many patients as possible, but keeping patients alive in poorer health states was viewed as a burden on society.

> *‘So obviously this 20 people will live […] within 10 years they’ll be not horribly* [ill] *but kind of like probably the majority of their time they will be dependent on someone to help them, probably dependent on taking pills treatment and not doing things that they would enjoy doing because they probably disabled or the health is compromised’* [P3]

#### Theme 6: Logic Driven Decisions

This theme is characterised by the use of objective decision rules. Participants used mathematical reasoning to differing extents. As mentioned, some identified the QALY maximising answer early on and continued with this logic throughout. Others used more varied decision rules and defaulted to the QALY maximising answer in more difficult scenarios, concurrent with results from Bryan et al (21). The quantification of life into ‘person-years’ served not only as a decision-making heuristic but as a psychological buffer against the difficulty of the scenario.

> *‘I’m kind of working out in maths ways now. The fact that 20 times 5 is 100 and 8 times 10 is 80. So, I’m thinking about it the amount of years that people will live altogether and not just one person if that made sense.’* [P6]

> *‘I think that’s why again I’m just going to go with 10 because that mathematically is the only sort of reason I can kind of come up with without any extra context’* [P8]

By using a QALY maximisation approach, participants were able to turn the decision into one of expressing the goodness of each outcome quantitatively and solving a mathematical equation. However, there were some cases where participants calculated the mathematically correct answer, and consciously chose to move away from this answer as even though this achieved an equal number of total QALYs, it still did not feel fair;

> *‘I went for the mathematical one last time, but only living for 6 months, I think the maths goes a bit out the window because year, my gut wants to go with that’* [P4]

This highlights an internal conflict between logic and emotion in respect to decisions around distributional equity. When the health gain was very small, fewer participants used mathematical reasoning, suggesting that extreme values (a large number of beneficiaries, a small health gain) elicited a more emotional reaction than more moderate scenarios. However, when participants were unable to make the decision, they sometimes reverted back to mathematical reasoning as a well-defined set of decision rules they were able to fall back on.

> *‘I think that with the health [score 60%] that’s just a bit too hard. So I’m going to go for the mathematically correct one. Yep.’* [P4]

For some participants, logic-driven decision making was an active and explicit choice; they actively suppressed any emotional response to maintain an arms-length decision-making role.

> *‘That’s why I’m basing it on number of years because you’re taking all of the humanity almost out of the question […] equal can only be equal if you take emotion out’* [P8]

Because the hypothetical patients lacked context, some participants attempted to fill in the blanks with imagined contexts, while others reacted by reducing the scenario to a mathematical exercise. This may be an example of rational compassion (25); filling in information gaps with real human characteristics leads participants to display empathy for the patients and irrational attachment to these identical and abstract stick figures. Actively avoiding these allowed participants to take a detached approach to find a solution that felt equal to them.

## Discussion

This study aimed to investigate how participants make trade-off decisions in a PTO task and, whilst research eliciting preferences for the concentration and dispersion of healthcare benefits exists (5,6,26), this study was the first to qualitatively investigate the reasoning behind such preferences. The findings concur closely with those of previous qualitative studies of PTOs, suggesting that although the specific trade-off presented to participants is distinct, the underlying reasoning processes used to navigate difficult healthcare decisions may be transferable across different decision-making contexts. For example, Damschroder et al (27). and De Silva et al. (28), alongside the current study, identified themes in which participants drew on their own experiences when making decisions, with participants in each study considering factors beyond the maximisation of total health gain. Equality of access to treatment was prioritised by participants in the current study and in De Silva et al. over the potential total health gains that could be achieved. Although not identified as a theme in De Silva et al., both the current study and Damschroder et al. also found that participants considered the wider societal consequences of their decisions. The consistency of these findings across different PTO tasks suggests that, when faced with complex healthcare trade-offs, participants may draw on a common set of underlying values and reasoning strategies, including personal experience, considerations of fairness and equality, and perceptions of wider societal impact. This suggests that qualitative investigation of PTOs can provide insights not only into preferences for the specific distributional trade-offs being examined, but also into the broader processes through which individuals construct and justify preferences when making difficult healthcare resource allocation decisions.

The themes highlight that the reasonings participants used generally fell into one of two schools of thought in their decision making. Firstly, those that transformed the survey into real-world scenarios, humanising the hypothetical participants with consideration of what they would be able to achieve in the time they have remaining, the level of suffering they would experience or the impact on society of keeping a certain characteristic of patient alive. On the other hand, some participants completely rejected this way of thinking and instead viewed the task as a purely quantitative exercise, in line with NICE’s current QALY maximisation approach. Participants showed emotional detachment from the context of the question and made decisions purely on the basis of equalising the total number of person-years allocated to each scenario. There was some support for S-shaped preferences: participants clearly expressed that health gains below 1-2 years was not valuable, and when presented with scenarios in the questions where the health gain was shorter (2 years or less), many remarked how the gain ‘wasn’t enough time’, or that the patients would not be able to achieve anything meaningful in this period.

From the qualitative results alone it is not possible to ascertain the shape of these preferences, many participants displayed a clear divergence away from QALY maximisation preferences. Many participants were strongly death averse, and displayed ‘rule of rescue’ (29) type reactions when faced with a scenario where participants would die should they not receive the treatment. Additionally, some participants highlighted that they attributed the value of a particular health gain to what was able to be achieved within this time frame, suggesting a potential convex curve at higher levels of health gain, and divergence away from previous studies that have studied concentration and dispersion preferences (5,6,26).

### Strengths and implications

This study was the first to conduct a qualitative investigation into S-Shaped preferences. The key strengths of this study lie in the pilot study and depth of the data. The pilot study allowed the PTO study to be adapted to ensure maximum understanding. A key message from this study is how public preferences in healthcare are more nuanced than is able to be captured through quantitative preference elicitation studies alone. Specifically, it can help explain the opposing results of previous studies: while Nord et al. (30) demonstrated that the public heavily favours egalitarian principles over QALY maximisation, Bryan et al. (31) showed that public preferences can largely conform to QALY-maximising assumptions. This highlights that preferences may depend on the level of emotional involvement of the participants, which may differ between survey designs. Repeated use of previous PTO designs would allow direct comparability of results and promoting the use of two stage quantitative and qualitative study designs would allow for greater understanding of distributional preferences.

### Limitations

Firstly, all participants were recruited through an online research panel, which may introduce self-selection bias toward digitally competent, ‘professional’ respondents who may be more detached and rational in their problem solving rather than the wider public in a real-world setting. This study is also subject to hypothetical bias, where some participants may have answered differently than in a real-world scenario. However, there is no revealed preference equivalent of this task from the public, as the decisions in choice tasks would almost certainly never be faced as a private citizen. Thus, the use of stated preference tasks is unavoidable.

There were several framing effects which this study did not examine, including varying the age of participants, and the pool size of the population. Previous research has shown that the pool size of potential patients does not alter participant responses to similar types of questions (5), other studies (32) show that this does have an impact. In the scenarios, participants were allocating health gains to a population of 200 people. If the participants were asked to make similar decisions relating to a population size more similar to the UK, responses and rationales might have been different as patients are “one of many” and the humanisation of the patients may have occurred to a lesser extent (33).

The task the participants were asked to complete was cognitively burdensome, evidenced by the use of heuristics by participants (23). However, the pilot showed high levels of understanding initially, which was only improved through iterative improvements of the survey design.

## Conclusion

In conclusion, whilst decision-making processes are heterogeneous, participants generally make decisions based on normative values, where decisions were guided by underlying values or rules about what ‘*ought to be’* (34).

The themes identified highlight that these values were heterogenous between participants, where some focused on the impact of the treatment to the patient, compared to societal impact. Some participants placed higher priority on the quality of life than others, and participants engaged or detached themselves from the question to varying degrees. The more detached patients displayed higher degrees of internal consistency and generally responded inline with a QALY maximisation framework. As such, stated preference tasks are useful tools, but if funding decisions are to reflect societal values, more data is needed on what shape these preferences take, and how they vary across the population. Further research should aim to understand the shape of preferences with respect to the concentration and dispersion of health gains to more closely align policy decision making processes with UK public preferences on distributional equity.

## Data Availability

Transcripts are available upon request.

